# Sleep Variability From Routine Nursing Observation and Wearable Actigraphy and Post-Enrollment Length of Stay in Acute Psychiatric Inpatients: Prospective Observational Study

**DOI:** 10.64898/2026.09.08.26362319

**Authors:** Hong-Ming Chen, Yi-Lung Chen, Cheryl C-H Yang

## Abstract

**Background:** Length of stay (LOS) in acute psychiatric inpatient settings is clinically and operationally important, but practical early markers for identifying patients at risk for prolonged hospitalization remain limited.

**Objective:** To evaluate whether sleep instability measured during the early inpatient observation period was associated with prolonged post-enrollment hospitalization in acute psychiatric inpatients.

**Methods:** This observational study included 94 consecutive patients admitted to an acute psychiatric inpatient unit. Monitoring began after study enrollment, and participants contributed 5 to 7 available nightly observations from wrist actigraphy and routine nursing observation; observations were not required to occur on consecutive calendar days. The primary exposure was intraindividual sleep variability (IISV), defined as the within-subject standard deviation of nightly total sleep time across available post-enrollment observations. The primary outcome was prolonged post-enrollment length of stay, defined using the 75th percentile of the cohort distribution. Age- and sex-adjusted logistic regression models evaluated associations, discrimination, calibration, and risk stratification. Total length of stay from admission was examined as a temporally distinct sensitivity estimand.

**Results:** Participants were enrolled a median of 4 days after admission (IQR 2-7) and contributed 5 to 7 available nightly observations, which accumulated over a median of 8 calendar days (IQR 7-10); 39 of 94 observation series (41.5%) fell on consecutive calendar days. Prolonged post-enrollment hospitalization was defined as more than 24.0 days, yielding 23 events among 94 participants. Greater actigraphy-derived IISV was associated with prolonged post-enrollment hospitalization (adjusted odds ratio [OR], 2.30 per 1-hour increase; 95% CI, 1.08-4.93; *P*=.03), with an area under the receiver operating characteristic curve (AUC) of 0.756 (95% CI, 0.626-0.852; optimism-corrected, 0.725) compared with 0.692 for a model containing age and sex alone. Nursing-derived IISV showed an association of similar direction (adjusted OR, 3.10; 95% CI, 1.15-8.39; *P*=.03; AUC, 0.723; 95% CI, 0.571-0.837; optimism-corrected, 0.690). A combined model containing both measures did not improve discrimination over actigraphy alone (AUC, 0.751).

**Conclusions:** Sleep instability measured during the early inpatient observation period, derived independently from wearable actigraphy and from routine nursing sleep documentation, was associated with prolonged post-enrollment hospitalization. Because the outcome interval overlaps the period during which the sleep observations were accumulated, these findings describe an association across the early inpatient course rather than a prediction made at enrollment. The results support further evaluation of repeated sleep variability as a longitudinal inpatient measure; the variable observation schedule, single-center sample, and absence of external validation mean that clinical implementation remains premature.

## Introduction

Acute psychiatric hospitalization is characterized by substantial heterogeneity in recovery trajectories and discharge timing. Length of stay (LOS) is clinically and operationally important, but the course of acute psychiatric illness unfolds over days rather than at a single assessment point. Practical measures that summarize this evolving inpatient course remain limited.[1,2]

Sleep disturbance is a transdiagnostic feature of psychiatric illness.[3] Most sleep assessments emphasize mean duration or measurements from individual nights, whereas intraindividual sleep variability (IISV) captures a different property: the extent to which sleep remains unstable across repeated observations. Such instability may reflect fluctuations in behavioral, circadian, or neurobiological regulation that are not represented by mean sleep duration alone.[3,4]

Wrist actigraphy permits repeated objective measurement of sleep-wake patterns, while routine nursing sleep observation is already embedded in acute psychiatric ward practice.[6–8] The two sources differ substantially in how sleep is measured, but both can generate repeated nightly observations. If a variability signal derived from routine nursing documentation behaves similarly to one derived from actigraphy, structured nursing data could provide a low-burden source of longitudinal information for inpatient clinical informatics.

We therefore examined whether IISV derived independently from wearable actigraphy and routine nursing sleep records during the early inpatient observation period was associated with prolonged post-enrollment hospitalization in a diagnostically heterogeneous acute psychiatric cohort. IISV was prespecified as the primary sleep construct. We evaluated association, discrimination, calibration, risk stratification, and prespecified threshold sensitivity, and examined total LOS from admission as a temporally distinct sensitivity estimand. Monitoring was designed to accumulate up to seven observations under routine ward conditions rather than to impose a fixed seven-calendar-day window.[9,10]

## Methods

This study was reported in accordance with the Strengthening the Reporting of Observational Studies in Epidemiology (STROBE) reporting guideline.[11]

### Study Design and Participants

This observational study included 94 consecutive patients admitted to an acute psychiatric inpatient unit in southern Taiwan between September 2019 and March 2023. Diagnostic categories included schizophrenia-spectrum disorders, bipolar mania, and depressive or anxiety-related disorders.

Eligible participants were aged 15 to 80 years and admitted to the acute psychiatric ward during the study period. Exclusion criteria included compulsory admission status, obvious withdrawal symptoms related to alcohol or other substances, clinically evident delirium, intellectual disability, or dementia. Participants were approached for enrollment when their psychiatric condition was considered sufficiently stable to permit informed consent, which occurred at a median of 4 days after admission (IQR 2-7).

### Sleep Assessment and Observation Schedule

Sleep was evaluated using wrist actigraphy and routine nursing observation, recorded concurrently. Objective sleep-wake patterns were assessed using the XA-5 wrist actigraph (iBAlab, National Yang-Ming Chiao Tung University, Taiwan), a triaxial accelerometry-based device developed through iterative hardware refinement within the same laboratory platform. The sleep-wake detection algorithm is architecturally consistent with that of the previously validated XA-2 platform, which demonstrated high agreement with polysomnography-derived sleep parameters in a prior validation study. A detailed description of the XA-5 hardware, signal processing approach, and algorithm validation background is provided in Multimedia Appendix 1. Nursing-based sleep estimates were collected according to routine inpatient clinical practice.

Monitoring began early after admission, once the participant had been enrolled. Each participant was scheduled to contribute up to seven nightly observations. **Observations were not necessarily obtained on consecutive calendar days**: they were collected as routine ward operations allowed, so the calendar period over which a participant’s observations accumulated varied between individuals. The number of available observations and the calendar span of each participant’s observation series are reported in Table 1 and are described in the Results.

Actigraphy and nursing observations were paired by night, so that each available nightly observation contributed a matched pair of estimates wherever both sources recorded that night.

### Sleep Variability

Intraindividual sleep variability was defined as the within-subject standard deviation (ddof = 1) of nightly total sleep time, computed across each participant’s available post-enrollment nightly observations. Because the number of available observations and the calendar span over which they were obtained varied between participants, this measure reflects night-to-night variability across the observations actually available for a given participant rather than across a fixed calendar window.

### Outcome Definition

The primary outcome was post-enrollment length of stay, defined as the interval from study enrollment to hospital discharge. Prolonged post-enrollment hospitalization was defined as a post-enrollment length of stay exceeding the 75th percentile of the cohort distribution, which corresponded to more than 24.0 days and identified 23 of 94 participants (24.5%). Sensitivity analyses used the corresponding 70th and 80th percentile thresholds, which corresponded to more than 23.0 days (26 events) and more than 29.0 days (16 events). Every participant remained at risk for the entire post-enrollment interval, and no participant was excluded. Total LOS, defined from hospital admission to discharge, was analyzed separately as a temporally distinct sensitivity estimand (75th percentile, more than 32.75 days; 24 events). Figure 1 shows the temporal relationship among study enrollment, observation acquisition, and the two length-of-stay estimands.

**Figure 1.**
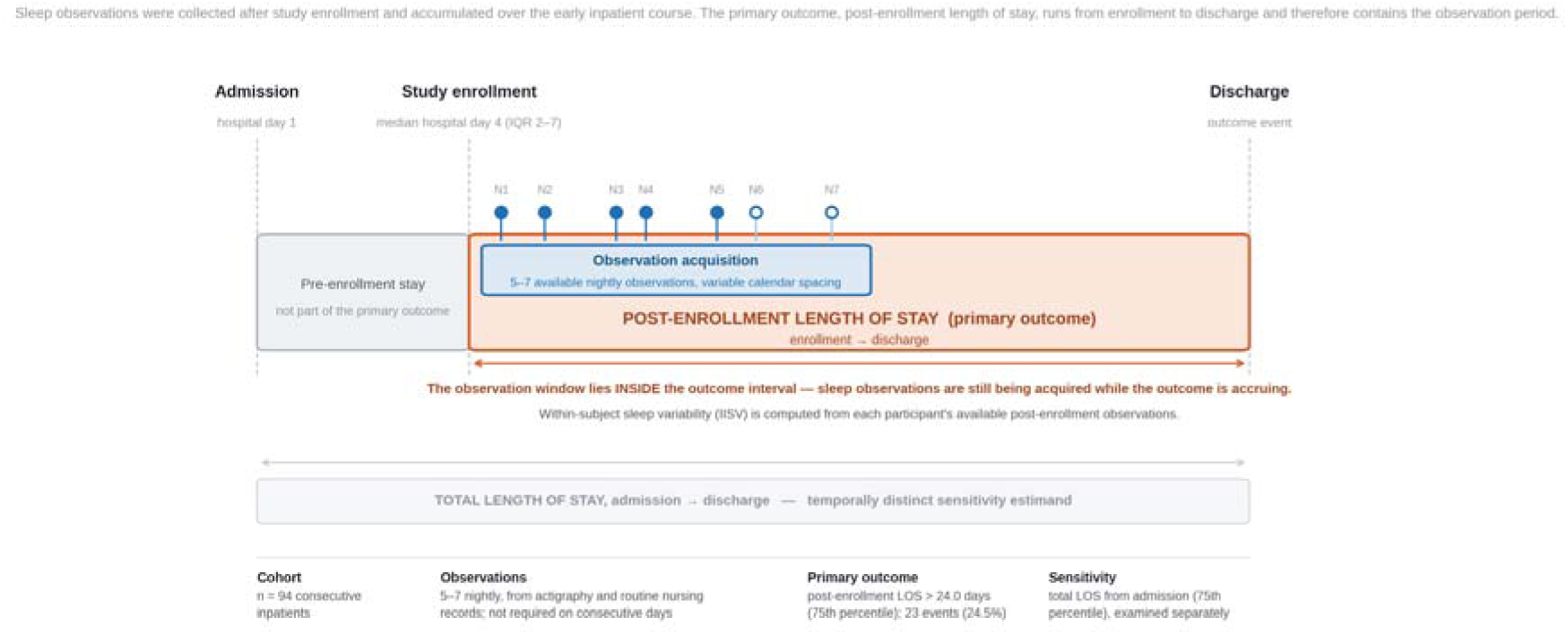
Study timing, sleep observations and outcome definitions. Sleep observations were collected after study enrollment and accumulated over the early inpatient course. The primary outcome, post-enrollment length of stay, runs from enrollment to discharge and therefore contains the observation period. Total length of stay from admission was examined as a temporally distinct sensitivity estimand. N1-N7 denote the nightly observations; participants contributed 5 to 7 available observations, which were not required to occur on consecutive calendar days.

### Statistical Analysis

Multivariable logistic regression models adjusted for age and sex were used to estimate associations between IISV and prolonged post-enrollment hospitalization. Odds ratios were calculated per 1-hour increase in sleep variability. Actigraphy-derived and nursing-derived IISV were examined in parallel, with a combined model used to assess whether the two measurement sources contributed overlapping or additive information.

Model discrimination was evaluated using the area under the receiver operating characteristic curve (AUC) with 95% confidence intervals and bootstrap optimism correction (Harrell bootstrap, 2000 replicates; eTable 7). Calibration was assessed by comparing predicted and observed probabilities across prespecified risk groupings, and risk stratification was summarized across tertiles of predicted probability. Sensitivity analyses examined alternative post-enrollment LOS thresholds and additional adjustment for diagnostic category. The same prespecified candidate sleep indices were re-evaluated as a robustness analysis; IISV remained the a priori primary construct and was not selected on the basis of this screen.

Total LOS from admission was examined as a separate temporal sensitivity estimand because it includes hospital days preceding study enrollment and therefore preceding acquisition of the repeated sleep observations.

This study was approved by the Institutional Review Board of Chang Gung Memorial Hospital, Taiwan (approval number: 2303260008) and was conducted in accordance with the Declaration of Helsinki. Written informed consent was obtained from all participants prior to any study procedures.

## Results

### Enrollment Timing and Observation Sampling Structure

The study included 94 psychiatric inpatients. Participants were enrolled a median of 4 days after admission (IQR 2-7; range 1-39), consistent with a monitoring period beginning early in the inpatient course.

Most participants contributed the full complement of seven available nightly observations: 86 of 94 (91.5%) for actigraphy and 84 of 94 (89.4%) for nursing observation, with the remainder contributing five or six. The median number of available observations was 7 (IQR 7-7, range 5-7) for both sources.

These observations were not obtained as a uniform block of consecutive calendar days. The observation series spanned a median of 8 calendar days (IQR 7-10, range 5-19), and only 39 of 94 series (41.5%) comprised observations on consecutive calendar days. Sleep variability estimates therefore summarize night-to-night variability across each participant’s available observations rather than across a fixed seven-day calendar window. The sampling structure is reported in full in Table 1.

### Baseline Characteristics

Baseline characteristics are summarized in Table 1. Mean age was 43.1 years (SD, 14.8), and 58 participants (61.7%) were male. Diagnostic categories included schizophrenia-spectrum disorders (32 patients [34.0%]), bipolar mania (31 patients [33.0%]), and depressive or anxiety-related disorders (31 patients [33.0%]). Mean actigraphy-derived total sleep time was 7.3 hours (SD, 1.2), and mean nursing-derived total sleep time was 7.1 hours (SD, 1.1). Mean actigraphy-derived sleep variability was 1.13 hours (SD, 0.66), and mean nursing-derived sleep variability was 1.14 hours (SD, 0.50). Median post-enrollment length of stay was 17.0 days (IQR, 12.0-24.0; range, 5-68). Mean total LOS from admission was 26.7 days (SD, 14.4), with a median of 23.0 days (IQR, 16.0-32.8).

### Actigraphy-Derived Sleep Variability and Model Performance

Greater actigraphy-derived IISV was associated with prolonged post-enrollment hospitalization (adjusted OR, 2.30 per 1-hour increase; 95% CI, 1.08-4.93; *P*=.03). Expressed per 1 SD of the index, the adjusted OR was 1.74 (95% CI, 1.05-2.89). In the same model, neither age (adjusted OR, 1.04 per year; 95% CI, 1.00-1.07; *P*=.05) nor male sex (adjusted OR, 2.56; 95% CI, 0.80-8.15; *P*=.11) reached conventional significance (Table 2).

The actigraphy-based model showed moderate discrimination (AUC, 0.756; 95% CI, 0.626-0.852; optimism-corrected AUC, 0.725), compared with an AUC of 0.692 for a model containing age and sex alone (Figure 2A). Calibration across quintiles of predicted risk showed agreement between predicted and observed probabilities with overlapping 95% confidence intervals (bootstrap-corrected calibration slope, 0.85; Brier score, 0.160; Hosmer-Lemeshow *P*=.89; Figure 2B and eTable 2).

**Figure 2.**
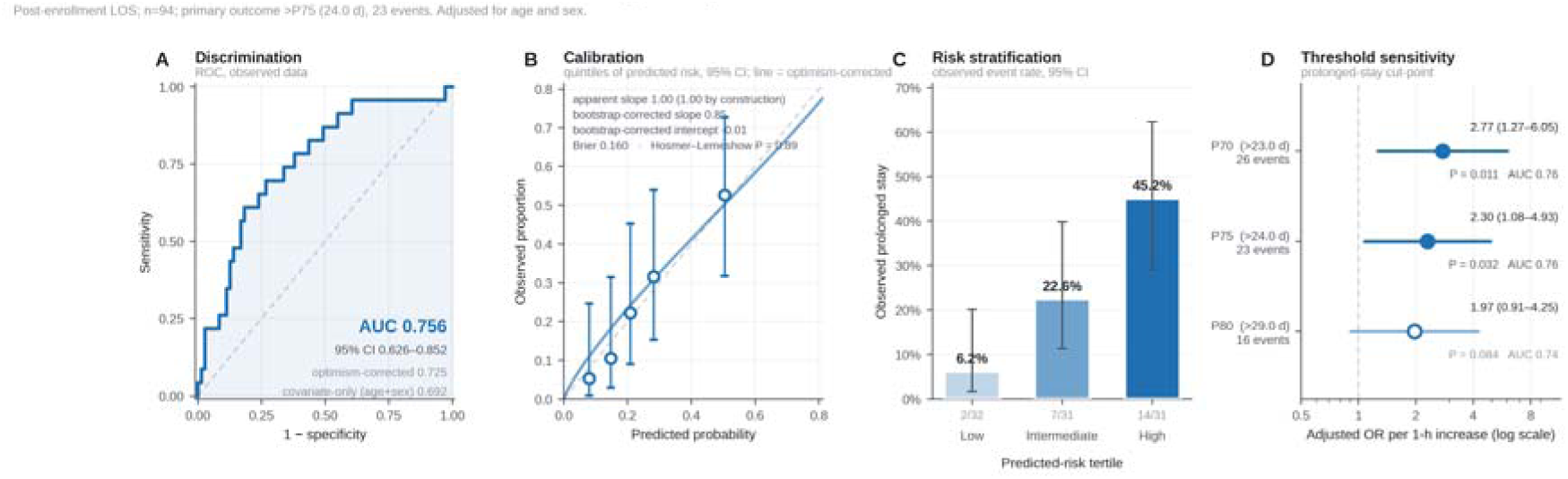
Actigraphy-derived sleep variability and post-enrollment length of stay (n=94; prolonged stay defined as more than 24.0 days, 23 events; models adjusted for age and sex). (A) Discrimination, shown as the receiver operating characteristic curve with the apparent and optimism-corrected area under the curve and the covariate-only reference. (B) Calibration across quintiles of predicted risk, with 95% confidence intervals; the fitted line is the bootstrap optimism-corrected calibration curve. (C) Observed rate of prolonged post-enrollment hospitalization by tertile of predicted risk, with 95% confidence intervals. (D) Sensitivity of the adjusted odds ratio to the prolonged-stay cut-point across the 70th, 75th and 80th percentiles.

Observed rates of prolonged post-enrollment hospitalization increased across tertiles of predicted risk, from 6.2% (2/32) in the lowest tertile to 22.6% (7/31) in the intermediate tertile and 45.2% (14/31) in the highest tertile (Figure 2C and eTable 3).

### Nursing-Derived Sleep Variability and Comparative Performance

Nursing-derived IISV showed an association of similar direction and magnitude (adjusted OR, 3.10 per 1-hour increase; 95% CI, 1.15-8.39; *P*=.03; OR per 1 SD, 1.76; 95% CI, 1.07-2.91). Discrimination was comparable to the actigraphy-based model (AUC, 0.723; 95% CI, 0.571-0.837; optimism-corrected AUC, 0.690; Figure 3A).

**Figure 3.**
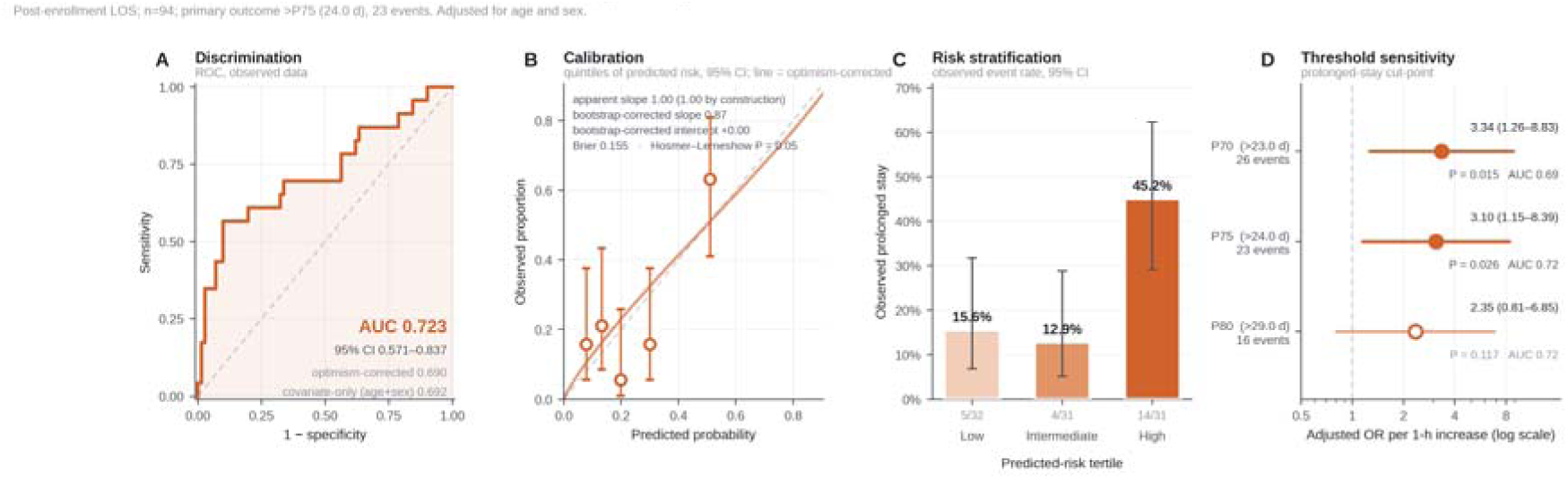
Nursing-derived sleep variability and post-enrollment length of stay (n=94; prolonged stay defined as more than 24.0 days, 23 events; models adjusted for age and sex). Panels A to D follow the same format as Figure 2.

Calibration of the nursing-derived model was less satisfactory than that of the actigraphy-based model (bootstrap-corrected calibration slope, 0.87; Brier score, 0.155; Hosmer-Lemeshow *P*=.047; Figure 3B), indicating some misfit between predicted and observed probabilities.

Risk stratification also differed between the two sources. Whereas the actigraphy-based model separated all three tertiles, the nursing-derived model separated only the highest tertile: observed rates were 15.6% (5/32) in the lowest tertile, 12.9% (4/31) in the intermediate tertile, and 45.2% (14/31) in the highest tertile (Figure 3C and eTable 3). The lowest and intermediate nursing-derived tertiles were therefore not distinguishable from one another.

In a combined model containing both actigraphy-derived and nursing-derived IISV, discrimination was not improved over the actigraphy-based model alone (AUC, 0.751; 95% CI, 0.608-0.854; optimism-corrected AUC, 0.702; eTable 6). This pattern is consistent with the two measurement sources carrying overlapping rather than additive information, although the present sample cannot resolve the extent of that overlap.

### Sensitivity Analyses

#### Alternative outcome thresholds

At the 70th percentile (more than 23.0 days; 26 events), the adjusted OR was 2.77 (95% CI, 1.27-6.05; *P*=.01; AUC, 0.765) for actigraphy-derived IISV and 3.34 (95% CI, 1.26-8.83; *P*=.02; AUC, 0.686) for nursing-derived IISV. At the 80th percentile (more than 29.0 days; 16 events), estimates were 1.97 (95% CI, 0.91-4.25; *P*=.08; AUC, 0.744) and 2.35 (95% CI, 0.81-6.85; *P*=.12; AUC, 0.717), respectively (eTable 4).

Associations were present at the 70th- and 75th-percentile definitions but were less precise and did not reach conventional significance at the 80th percentile, where only 16 events were available.

#### Candidate-index robustness screen

The 16 prespecified candidate sleep indices were each entered into the identical age- and sex-adjusted model (eFigure 1 and eTable 1).

Actigraphy-derived IISV ranked first of 16 by adjusted AUC (0.756), with the largest increment over the age- and sex-only model (ΔAUC, +0.065). IISV was prespecified a priori on construct grounds and was not selected from the candidate-index screen; its ranking is reported for transparency. No candidate index survived Benjamini-Hochberg correction across the 16 comparisons (minimum q=.15). This screen should therefore be read as a robustness analysis rather than as evidence that IISV is a statistically selected optimal index.

#### Diagnosis-adjusted models

After additional adjustment for diagnostic category, the association with actigraphy-derived IISV was attenuated (adjusted OR, 2.05; 95% CI, 0.92-4.56; *P*=.08), whereas the association with nursing-derived IISV remained (adjusted OR, 3.11; 95% CI, 1.13-8.59; *P*=.03). Diagnostic category itself was not independently associated with the outcome in either model (schizophrenia-spectrum versus depressive or anxiety-related disorders: adjusted OR, 2.15; 95% CI, 0.55-8.38; *P*=.27 in the actigraphy model and 2.87; 95% CI, 0.75-10.95; *P*=.12 in the nursing model) (eTable 5).

#### Total LOS from admission

Under the temporally distinct total-LOS estimand (more than 32.75 days; 24 events), associations were weaker and did not reach conventional significance for either source: actigraphy-derived IISV, adjusted OR 1.87 (95% CI, 0.91-3.85; *P*=.09; AUC, 0.711; optimism-corrected, 0.676); nursing-derived IISV, adjusted OR 1.92 (95% CI, 0.75-4.93; *P*=.18; AUC, 0.709; optimism-corrected, 0.673). Total LOS includes hospital days that preceded enrollment and therefore preceded acquisition of the repeated sleep observations (eTable 8).

## Discussion

### Principal Results

In this cohort of acute psychiatric inpatients, greater early-course sleep variability was associated with prolonged post-enrollment hospitalization when variability was derived independently from wearable actigraphy (adjusted OR, 2.30; 95% CI, 1.08-4.93) and from routine nursing observations (adjusted OR, 3.10; 95% CI, 1.15-8.39). Discrimination was moderate for both sources (AUC, 0.756 and 0.723; optimism-corrected, 0.725 and 0.690) against an age- and sex-only model of 0.692, and a combined model did not improve on actigraphy alone. The convergence of these two measurement streams is notable because they capture sleep through different processes. The finding therefore points less to a particular device than to sleep instability as a potentially informative feature of the inpatient course.

Unlike static sleep measures, IISV summarizes fluctuation across repeated observations. This may capture an aspect of acute-state instability that is obscured by mean sleep duration alone. The present observational data cannot establish the biological mechanism underlying this association, and the modest sample limits precision, but the parallel direction of findings across actigraphy and nursing records supports further evaluation of variability-based sleep measures.[3–5]

The temporal definition of LOS is important. Post-enrollment LOS begins at study enrollment and therefore covers the interval during which repeated sleep observations were accumulated. Total LOS from admission additionally incorporates hospital days that occurred before enrollment and before the sleep series was acquired. These estimands answer related but different questions; the attenuation observed under the total-LOS sensitivity analysis should therefore be interpreted as sensitivity to the temporal origin of the outcome rather than as a simple replication failure.

An important feature of the study is that monitoring reflected routine acute-ward conditions rather than a protocol-fixed calendar window. Participants contributed 5 to 7 available observations, and these observations were not necessarily consecutive. IISV should therefore be interpreted as variability across the available early inpatient observations, not as variability during a uniform first seven calendar days. Any future clinical implementation would need to specify the acquisition schedule explicitly.

Routine nursing observation is particularly relevant because it is already embedded in psychiatric inpatient care. If routinely documented sleep information can be captured in structured, computable form, variability measures could be evaluated without requiring additional monitoring equipment. Routine nursing observations and actigraphy appear to encode overlapping longitudinal information, and combined modeling did not improve discrimination over actigraphy alone. The present comparison should not be interpreted as evidence that actigraphy replaces nursing assessment, nor that the two sources are interchangeable; the nursing-derived model showed weaker calibration and separated only its highest risk tertile, so the two streams behave similarly in overall discrimination but not in how finely they rank individual participants.[7,8,12]

### Limitations

Several limitations should be considered. First, this was a single-center study with a modest sample size, limiting precision and external generalizability. Second, the observational design does not permit causal inference. Third, enrollment required sufficient clinical stability for informed consent and occurred after admission, which may have introduced selection related to early clinical course.

Fourth, the observation schedule was not uniform. Participants contributed 5 to 7 available nightly observations over variable calendar spans, and observations were not always obtained on consecutive days. Reasons for missed or delayed observations were not systematically recorded, so informative observation scheduling cannot be excluded. Two participants had usable actigraphy observations without verifiable nightly timestamps, and several additional actigraphy observations lacked date information; these data contributed to IISV but limit complete reconstruction of acquisition timing.

Fifth, post-enrollment LOS overlaps temporally with the period during which sleep observations were accumulated. The analysis therefore evaluates an association between early-course sleep variability and the duration of the post-enrollment hospitalization; it should not be interpreted as a prediction made at enrollment or at a fixed landmark before all predictor observations were available.

Sixth, calibration differed between the two measurement sources. The nursing-derived model showed evidence of misfit between predicted and observed probabilities (Hosmer-Lemeshow *P*=.047) and did not separate its lowest from its intermediate risk tertile, so its risk estimates should be treated more cautiously than those of the actigraphy-based model.

Seventh, the cohort was diagnostically heterogeneous, and diagnosis-adjusted estimates should be interpreted cautiously given the sample size. Finally, the models were internally evaluated only; external validation in larger multicenter cohorts with prospectively specified observation schedules is required before clinical use.

### Comparison With Prior Work

These findings sit alongside a growing body of evidence linking actigraphy-derived sleep variability to clinically meaningful psychiatric outcomes. Prior work has established this association primarily in outpatient and community settings, with a focus on longer-term illness trajectories. A recent multicenter cohort study demonstrated that actigraphy-derived sleep phase variability and rest-activity amplitude predicted depressive relapse over 1 to 2 years of follow-up in adults with remitted major depressive disorder.[5] The present study asks a different question in a different setting: whether sleep instability observed during the early inpatient course is associated with the duration of the subsequent hospital course. The direction and magnitude of the present association are consistent with that literature, although the single-center sample and the overlap between the observation period and the outcome interval limit how far the comparison can be taken.

The parallel performance of nursing-derived sleep variability also relates to prior work comparing observational and device-based sleep assessment in psychiatric inpatients. Previous studies have demonstrated that nurse-rated sleep observations provide clinically relevant information that may complement actigraphy-derived data,[7,8] but the association of variability — rather than mean sleep duration — derived from routine nursing documentation with inpatient course has not been directly examined. The present findings suggest that night-to-night variability in nursing-documented sleep behaves similarly to its actigraphy-derived counterpart in overall discrimination, which is of interest for the informatics value of structured nursing sleep documentation.

### Conclusions

Sleep instability measured during the early inpatient observation period, derived independently from wearable actigraphy and from routine nursing sleep documentation, was associated with prolonged post-enrollment hospitalization in acute psychiatric inpatients. Because the outcome interval overlaps the period over which the sleep observations were accumulated, these results describe an association across the early inpatient course rather than a prediction made at enrollment or at a fixed landmark. The findings support further study of repeated sleep variability as a longitudinal inpatient measure, particularly where it can be derived from documentation already generated in routine care. The variable observation schedule, the single-center sample, and the absence of external validation preclude clinical implementation at present.

## Acknowledgments

The authors thank Chang Gung Memorial Hospital, Chiayi, Taiwan for institutional support. We are grateful to the nursing staff of the acute psychiatric ward for their dedication to patient care and for their diligent sleep documentation, which formed a central component of this study. We also thank the attending psychiatrists for their clinical collaboration and support throughout data collection. We extend our appreciation to the university for academic support and to the K & Y Lab for their contributions to the research environment. We are particularly grateful to Professor Po-Chao Kuo for developing the actigraphy device used in this study.

## Author Contributions

HMC conceived and designed the study, obtained funding, coordinated data collection, performed data analysis, interpreted the results, and drafted the manuscript. YLC contributed to participant recruitment and data collection and critically reviewed the manuscript. CCHY supervised the study, provided methodological and conceptual guidance, and critically revised the manuscript for important intellectual content. All authors read and approved the final manuscript.

## Data Availability

The data that support the findings of this study are available from the corresponding author upon reasonable request. Data sharing is subject to applicable deidentification requirements and general ethical principles governing the use of human participant data, in accordance with institutional review board oversight.

## Abbreviations

AUC: area under the receiver operating characteristic curve; CI: confidence interval; EHR: electronic health record; IQR: interquartile range; LOS: length of stay; OR: odds ratio; ROC: receiver operating characteristic; SD: standard deviation; STROBE: Strengthening the Reporting of Observational Studies in Epidemiology; TST: total sleep time

## Multimedia Appendix

**Multimedia Appendix 1.** Actigraphy Device Description and Validation Background (XA-5 Wrist Actigraph, iBAlab, National Yang-Ming Chiao Tung University, Taiwan). [DOCX File, uploaded separately]

## References

1. Horne CM, Hay K, Watson S, Anderson KN. An evaluation of sleep disturbance on in-patient psychiatric units. BJPsych Bull. 2018;42(5):193–197. doi:10.1192/bjb.2018.42

2. Bowers L, Nijman H, Allan T, Simpson A, Warren J, Turner L. Prevention and management of aggression training and violent incidents on U.K. acute psychiatric wards. Psychiatr Serv. 2006;57(7):1022–1026. doi:10.1176/ps.2006.57.7.1022

3. Wulff K, Gatti S, Wettstein JG, Foster RG. Sleep and circadian rhythm disruption in psychiatric and neurodegenerative disease. Nat Rev Neurosci. 2010;11(8):589–599. doi:10.1038/nrn2868

4. Fang Y, Forger DB, Frank E, Sen S, Goldstein C. Day-to-day variability in sleep parameters and depression risk: a prospective cohort study of training physicians. npj Digit Med. 2021;4:28. doi:10.1038/s41746-021-00400-z

5. Tonon AC, Palagini L, Geoffroy PA, et al. One-year actigraphy study of sleep and rest-activity rhythms as markers of relapse in depression. JAMA Psychiatry. 2026;83(4):379–388. doi:10.1001/jamapsychiatry.2025.2638

6. Conley S, Knies A, Batten J, et al. Agreement between actigraphic and polysomnographic measures of sleep in adults: a clinical review. Sleep Med Rev. 2019;46:151–160. doi:10.1016/j.smrv.2019.04.012

7. Krahn LE, Lin SC, Wisbey JA, O’Connor MK. Assessing sleep in psychiatric inpatients: nurse and patient reports versus wrist actigraphy. Ann Clin Psychiatry. 1997;9(4):203–210.

8. Richards KC, O’Sullivan PS, Phillips RL. Measurement of sleep in critically ill patients. J Nurs Meas. 2000;8(2):131–144. doi:10.1891/1061-3749.8.2.131

9. Aili K, Åström-Paulsson S, Stoetzer U, Svartengren M, Hillert L. Reliability of actigraphy and subjective sleep measurements in adults: the design of sleep assessments. J Clin Sleep Med. 2017;13(1):39–47. doi:10.5664/jcsm.6392

10. Lau T, Barger LK, Lockley SW, et al. Minimum number of nights for reliable estimation of habitual sleep using a consumer sleep tracker. npj Digit Med. 2022;5:40. doi:10.1038/s41746-022-00583-x

11. von Elm E, Altman DG, Egger M, Pocock SJ, Gøtzsche PC, Vandenbroucke JP; STROBE Initiative. The Strengthening the Reporting of Observational Studies in Epidemiology (STROBE) statement: guidelines for reporting observational studies. Ann Intern Med. 2007;147(8):573–577. doi:10.7326/0003-4819-147-8-200710160-00010

12. Mykkänen M, Kinnunen UM, Liljamo P, Ahonen O, Kuusisto A, Saranto K. Using standardized nursing data for knowledge generation: ward level analysis of point of care nursing documentation. Int J Med Inform. 2022;167:104879. doi:10.1016/j.ijmedinf.2022.104879

