## Supplementary Material for "Sleep Variability From Routine Nursing Observation and Wearable Actigraphy and Post-Enrollment Length of Stay in Acute Psychiatric Inpatients: Prospective Observational Study"

All supplementary analyses use the primary estimand: post-enrollment length of stay, measured from study enrollment to discharge, in the full cohort of 94 participants. Prolonged post-enrollment hospitalization is defined as more than 24.0 days (75th percentile of the cohort distribution), giving 23 events. Unless stated otherwise, models are logistic and adjusted for age and sex, odds ratios are per 1-hour increase in intraindividual sleep variability, and optimism correction uses a Harrell bootstrap with 2000 replicates (seed 20260915). eTable 8 reports the total length of stay from admission as a temporally distinct sensitivity estimand.

**eTable 1. Prespecified candidate sleep indices and prolonged post-enrollment length of stay**

| **Rank by AUC** | **Sleep index** | **Source** | **Domain** | **n** | **Events** | **Adjusted OR per 1 SD (95% CI)** | **P** | **FDR q** | **Model AUC (95% CI)** | **ΔAUC vs covariate-only** | **DeLong P** |
| --- | --- | --- | --- | --- | --- | --- | --- | --- | --- | --- | --- |
| 1 | Actigraphy TST variability (within-subject SD), h | wearable | variability | 94 | 23 | 1.74 (1.05–2.89) | .032 | 0.15 | 0.756 (0.626–0.852) | +0.065 | .079 |
| 2 | Actigraphy WASO variability (SD), h | wearable | variability | 94 | 23 | 1.60 (0.98–2.62) | .061 | 0.15 | 0.745 (0.611–0.844) | +0.053 | .182 |
| 3 | Actigraphy wake after sleep onset (mean), h | wearable | level | 94 | 23 | 1.83 (1.12–2.98) | .016 | 0.15 | 0.738 (0.605–0.838) | +0.046 | .295 |
| 4 | Actigraphy TST coefficient of variation | wearable | variability | 94 | 23 | 1.52 (0.94–2.46) | .087 | 0.17 | 0.733 (0.601–0.834) | +0.041 | .188 |
| 5 | Actigraphy time in bed variability (SD), h | wearable | variability | 94 | 23 | 1.37 (0.82–2.29) | .223 | 0.32 | 0.726 (0.596–0.827) | +0.035 | .170 |
| 6 | Actigraphy awakenings variability (SD), n | wearable | variability | 94 | 23 | 1.57 (0.97–2.55) | .066 | 0.15 | 0.725 (0.602–0.821) | +0.033 | .381 |
| 7 | Actigraphy time in bed (mean), h | wearable | level | 94 | 23 | 1.64 (0.96–2.78) | .068 | 0.15 | 0.724 (0.586–0.829) | +0.032 | .376 |
| 8 | Nursing-recorded TST variability (within-subject SD), h | nursing | variability | 94 | 23 | 1.76 (1.07–2.91) | .026 | 0.15 | 0.723 (0.571–0.837) | +0.032 | .449 |
| 9 | Nursing-recorded TST coefficient of variation | nursing | variability | 94 | 23 | 1.51 (0.93–2.46) | .094 | 0.17 | 0.721 (0.568–0.835) | +0.029 | .401 |
| 10 | Actigraphy sleep efficiency (mean), % | wearable | level | 94 | 23 | 0.64 (0.40–1.03) | .064 | 0.15 | 0.716 (0.581–0.822) | +0.025 | .512 |
| 11 | Actigraphy awakenings per night (mean), n | wearable | level | 94 | 23 | 1.51 (0.91–2.51) | .113 | 0.18 | 0.715 (0.589–0.814) | +0.023 | .559 |
| 12 | Actigraphy total sleep time (mean), h | wearable | level | 94 | 23 | 1.06 (0.64–1.75) | .829 | 0.83 | 0.701 (0.561–0.811) | +0.009 | .142 |
| 13 | Actigraphy sleep efficiency variability (SD), % | wearable | variability | 94 | 23 | 1.25 (0.78–2.01) | .357 | 0.48 | 0.701 (0.563–0.811) | +0.009 | .580 |
| 14 | Nursing-recorded sleep interruptions (mean), n | nursing | level | 94 | 23 | 1.23 (0.77–1.97) | .394 | 0.48 | 0.699 (0.561–0.809) | +0.008 | .653 |
| 15 | Nursing-recorded interruptions variability (SD), n | nursing | variability | 94 | 23 | 0.92 (0.56–1.51) | .732 | 0.81 | 0.696 (0.561–0.805) | +0.005 | .653 |
| 16 | Nursing-recorded total sleep time (mean), h | nursing | level | 94 | 23 | 0.92 (0.56–1.52) | .755 | 0.81 | 0.694 (0.555–0.805) | +0.002 | .817 |

Each of the 16 prespecified candidate sleep indices was entered into the identical age- and sex-adjusted logistic model on the E2 cohort (n=94; prolonged post-enrollment length of stay defined as more than 24.0 days; 23 events). Odds ratios are expressed per 1 SD of the index so that indices on different scales are comparable. q = Benjamini-Hochberg false discovery rate across the 16 comparisons; no index survives correction (minimum q=.15). Intraindividual sleep variability was prespecified a priori on construct grounds and was not selected from this screen; its rank is reported for transparency only. Covariate-only (age + sex) model AUC 0.692.

**eTable 2. Calibration across quintiles of predicted risk**

| **Model** | **Quintile** | **n** | **Mean predicted probability** | **Observed events** | **Observed proportion** |
| --- | --- | --- | --- | --- | --- |
| Actigraphy-derived model | Q1 (lowest) | 19 | 0.080 | 1 | 5.3% |
|  | Q2 | 19 | 0.147 | 2 | 10.5% |
|  | Q3 | 18 | 0.209 | 4 | 22.2% |
|  | Q4 | 19 | 0.282 | 6 | 31.6% |
|  | Q5 (highest) | 19 | 0.504 | 10 | 52.6% |
| Nursing-derived model | Q1 (lowest) | 19 | 0.078 | 3 | 15.8% |
|  | Q2 | 19 | 0.133 | 4 | 21.1% |
|  | Q3 | 18 | 0.199 | 1 | 5.6% |
|  | Q4 | 19 | 0.301 | 3 | 15.8% |
|  | Q5 (highest) | 19 | 0.510 | 12 | 63.2% |

Participants ranked by predicted probability from the corresponding primary model and divided into quintiles. Hosmer-Lemeshow P=.89 for the actigraphy-derived model and P=.047 for the nursing-derived model.

**eTable 3. Risk stratification by predicted-risk tertile**

| **Model** | **Tertile** | **n** | **Mean predicted probability** | **Observed events** | **Observed proportion** |
| --- | --- | --- | --- | --- | --- |
| Actigraphy-derived model | Low | 32 | 0.102 | 2 | 6.2% |
|  | Intermediate | 31 | 0.211 | 7 | 22.6% |
|  | High | 31 | 0.426 | 14 | 45.2% |
| Nursing-derived model | Low | 32 | 0.097 | 5 | 15.6% |
|  | Intermediate | 31 | 0.203 | 4 | 12.9% |
|  | High | 31 | 0.438 | 14 | 45.2% |

Participants ranked by predicted probability and divided into tertiles (32/31/31). The actigraphy-derived model separates all three tertiles; the nursing-derived model separates only the highest tertile, its lowest and intermediate tertiles being indistinguishable.

**eTable 4. Sensitivity across alternative prolonged-stay thresholds**

| **Threshold** | **Cut-point, d** | **Events** | **Event rate** | **Actigraphy OR (95% CI)** | **P** | **AUC** | **Nursing OR (95% CI)** | **P** | **AUC** |
| --- | --- | --- | --- | --- | --- | --- | --- | --- | --- |
| P70 | >23.0 | 26 | 27.7% | 2.77 (1.27–6.05) | .011 | 0.765 | 3.34 (1.26–8.83) | .015 | 0.686 |
| P75 (primary) | >24.0 | 23 | 24.5% | 2.30 (1.08–4.93) | .032 | 0.756 | 3.10 (1.15–8.39) | .026 | 0.723 |
| P80 | >29.0 | 16 | 17.0% | 1.97 (0.91–4.25) | .084 | 0.744 | 2.35 (0.81–6.85) | .117 | 0.717 |

Odds ratios per 1-hour increase in intraindividual sleep variability, adjusted for age and sex. Cut-points and event counts are recomputed at each percentile within the n=94 cohort. The 75th-percentile row is the primary analysis and remains primary irrespective of the other rows.

**eTable 5. Diagnosis-adjusted sensitivity analysis**

| **Model** | **Predictor** | **Adjusted OR (95% CI)** | **P value** |
| --- | --- | --- | --- |
| Actigraphy-derived model | Actigraphy-derived sleep variability (IISV), per 1 h | 2.05 (0.92–4.56) | .079 |
|  | Age, per 1 y | 1.04 (1.00–1.08) | .039 |
|  | Male sex | 2.17 (0.66–7.18) | .203 |
|  | Schizophrenia-spectrum vs depressive/anxiety | 2.15 (0.55–8.38) | .269 |
|  | Bipolar mania vs depressive/anxiety | 1.33 (0.34–5.17) | .680 |
|  | Model AUC (95% CI) | 0.767 (0.634–0.862) |  |
|  | Optimism-corrected AUC | 0.707 |  |
| Nursing-derived model | Nursing-derived sleep variability (IISV), per 1 h | 3.11 (1.13–8.59) | .029 |
|  | Age, per 1 y | 1.04 (1.00–1.08) | .060 |
|  | Male sex | 1.65 (0.52–5.20) | .395 |
|  | Schizophrenia-spectrum vs depressive/anxiety | 2.87 (0.75–10.95) | .122 |
|  | Bipolar mania vs depressive/anxiety | 1.26 (0.31–5.04) | .746 |
|  | Model AUC (95% CI) | 0.748 (0.601–0.854) |  |
|  | Optimism-corrected AUC | 0.688 |  |

Models additionally adjusted for diagnostic category, with depressive or anxiety-related disorders as the reference. No interaction or subgroup analysis was performed.

**eTable 6. Actigraphy-derived, nursing-derived and combined models**

| **Model** | **n** | **Events** | **Actigraphy IISV OR (95% CI)** | **Nursing IISV OR (95% CI)** | **AUC (95% CI)** | **Optimism-corrected AUC** | **ΔAUC vs covariate-only** | **DeLong P** |
| --- | --- | --- | --- | --- | --- | --- | --- | --- |
| Covariate-only (age + sex) | 94 | 23 | — | — | 0.692 (0.552–0.803) | 0.670 | — | — |
| Actigraphy IISV | 94 | 23 | 2.30 (1.08–4.93) | — | 0.756 (0.626–0.852) | 0.725 | +0.065 | .079 |
| Nursing IISV | 94 | 23 | — | 3.10 (1.15–8.39) | 0.723 (0.571–0.837) | 0.690 | +0.032 | .449 |
| Combined (actigraphy + nursing IISV) | 94 | 23 | 1.74 (0.75–4.02) | 2.20 (0.71–6.77) | 0.751 (0.608–0.854) | 0.702 | +0.059 | .167 |

All models additionally adjusted for age and sex and fitted on the same 94 participants. DeLong P values compare each model with the covariate-only model.

**eTable 7. Internal validation by bootstrap optimism correction**

| **Model** | **Index** | **Apparent** | **Optimism** | **Optimism-corrected** | **Replicates** | **Seed** |
| --- | --- | --- | --- | --- | --- | --- |
| Actigraphy-derived model | AUC | 0.7563 | +0.0316 | 0.7247 | 1996 of 2000 | 20260915 |
|  | Calibration slope | 1.0000 | +0.1468 | 0.8532 | 1996 of 2000 | 20260915 |
|  | Calibration intercept | -0.0000 | +0.0075 | -0.0075 | 1996 of 2000 | 20260915 |
|  | Brier score (apparent) | 0.1598 | — | — | 1996 of 2000 | 20260915 |
| Nursing-derived model | AUC | 0.7232 | +0.0328 | 0.6904 | 1996 of 2000 | 20260915 |
|  | Calibration slope | 1.0000 | +0.1261 | 0.8739 | 1996 of 2000 | 20260915 |
|  | Calibration intercept | -0.0000 | -0.0031 | 0.0031 | 1996 of 2000 | 20260915 |
|  | Brier score (apparent) | 0.1549 | — | — | 1996 of 2000 | 20260915 |

Harrell bootstrap optimism correction. The full model-fitting procedure was repeated in every replicate. The apparent calibration slope is 1.00 by construction for a model evaluated on its own fitting sample; the optimism-corrected slope is the informative quantity.

**eTable 8. Total length of stay from admission — temporal sensitivity estimand**

| **Measurement source** | **n** | **Events** | **Cut-point, d** | **Adjusted OR (95% CI)** | **P value** | **AUC (95% CI)** | **Optimism-corrected AUC** |
| --- | --- | --- | --- | --- | --- | --- | --- |
| Actigraphy-derived sleep variability | 94 | 24 | >32.75 | 1.87 (0.91–3.85) | .090 | 0.711 (0.578–0.815) | 0.676 |
| Nursing-derived sleep variability | 94 | 24 | >32.75 | 1.92 (0.75–4.93) | .176 | 0.709 (0.567–0.819) | 0.673 |

Total length of stay measured from hospital admission, analysed as a temporally distinct sensitivity estimand using the identical model specification. Total length of stay includes hospital days that preceded study enrollment and therefore preceded acquisition of the repeated sleep observations; the weaker associations should be read as sensitivity to the temporal origin of the outcome rather than as a failed replication. These values are carried forward unchanged from the frozen total-LOS analysis and were not re-estimated.

**eFigure 1. Prespecified candidate sleep indices and post-enrollment length of stay**

Each of the 16 prespecified candidate sleep indices entered the identical age- and sex-adjusted logistic model on the E2 cohort (n=94; prolonged post-enrollment length of stay defined as more than 24.0 days; 23 events); indices are ordered by adjusted AUC. (A) Adjusted odds ratio per 1 SD of each index, with 95% confidence intervals; P and q columns give the unadjusted P value and the Benjamini-Hochberg false discovery rate across the 16 comparisons. (B) Model AUC with 95% confidence intervals; the dashed line marks the covariate-only (age and sex) model. The shaded row is intraindividual sleep variability, which was prespecified a priori on construct grounds and was not selected from this screen.

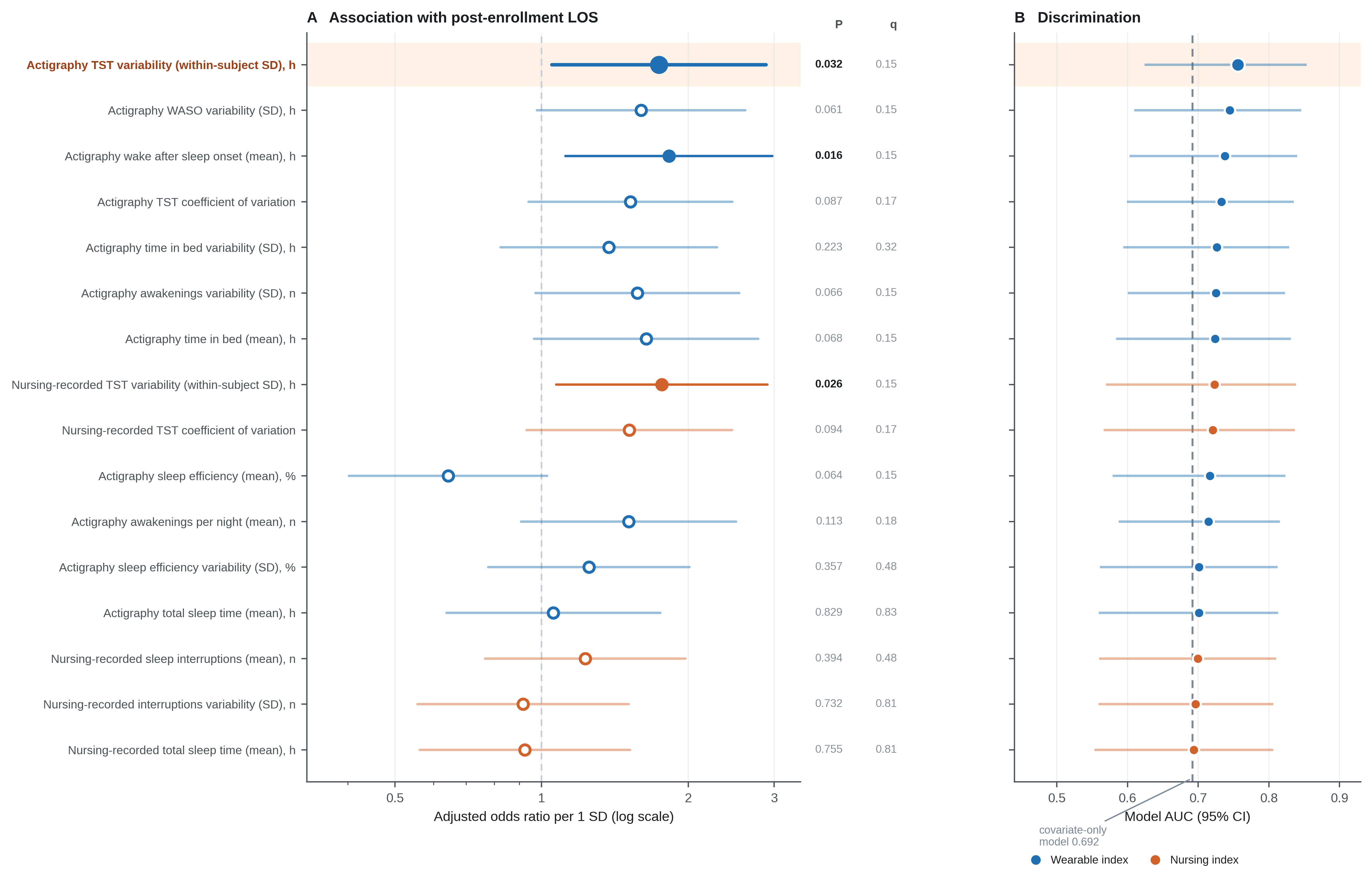
